# Study Protocol: Psychometric Testing of the German Vestibular Schwannoma Quality of Life Index - A Multicenter Study on Quality of Life and Patient-Centered Care in Vestibular Schwannoma

**DOI:** 10.1101/2025.07.08.25331128

**Authors:** Mareike Rutenkröger, Lasse Dührsen, Maximilian Scheer, Jannik Walter, Andrea Baehr, Bastian Baselt, Alexander Huber, Isabelle Scholl

## Abstract

**Introduction:** Vestibular schwannomas (VSs) are benign tumors of the vestibulocochlear nerve that often cause significant neurological and functional impairment, affecting patients’ overall quality of life (QoL). While clinical assessments have traditionally focused on hearing preservation and tumor control, patients often emphasize other critical symptoms such as dizziness, pain, cognitive difficulties and satisfaction with care. Therefore, patient-centered care that addresses the full range of patient experiences is essential. Despite its importance, patientcentered care in VS remains underexplored. This study will address this gap by psychometrically validating the German version of the Vestibular Schwannoma Quality of Life (VSQOL) Index, a newly developed QoL tool that includes key patient-centered domains. The primary objective is to validate the reliability and validity of the German VSQOL Index. The secondary aim is to assess VS patients’ experience of patient-centered care and its impact on their well-being.

**Methods and analysis:** This multicenter, cross-sectional study will involve German-speaking VS patients from several clinical centers in Germany and Switzerland as well as an online cohort. Psychometric testing of the German VSQOL will include reliability assessments (e.g. Cronbach’s alpha, test-retest reliability), confirmatory factor analysis and convergent validity. In parallel, the study will assess patient-centered experiences of care using the EPAT questionnaire.

**Discussion:** Ethical approval has been obtained and all participants will be asked to provide written informed consent. The results will be shared through scientific publications and conferences, as well as with patient groups, in order to support improvements in clinical care. By validating the German version of the VSQOL Index, this study will provide clinicians with a reliable tool for capturing patient-reported outcomes in VS, including symptoms that are often overlooked in standard assessments. Using the EPAT questionnaire alongside the VSQOL Index will provide insight into the relationship between patient experiences of care and quality of life. Together, these findings will support the delivery of more patient-centred and needs-oriented care in clinical practice and future research.

## Introduction

Vestibular schwannomas (VSs) are non-cancerous tumors of the vestibulocochlear nerve that often result in significant neurological and functional challenges. This impacts a patient’s overall well-being, whether due to disease progression or the effects of treatment (1). Studies comparing surgery and radiation therapy for VS show minimal differences in long-term quality of life (QOL), with neither treatment significantly altering QOL across most domains(2–4). While surgery may reduce anxiety due to the perception of tumor removal as a “cure,” the impact of the diagnosis itself on QOL often outweighs the effects of the treatment choice, highlighting the need for personalized patient counseling (2).

Given the complex and multifaceted impact of VS on patients, the importance of patientcentered care cannot be overstated. Patient-centered care emphasizes understanding and addressing the full range of patient experiences, preferences, and needs, particularly for chronic conditions such as VS, where QOL can be significantly affected by both the condition and its treatment (1,5). Despite the importance of this approach, there is a notable lack of studies that comprehensively explore the patient-centeredness of healthcare delivery in VS. Traditional clinical assessments typically focus on facial nerve function, hearing preservation, and tumor management (6–8). However, patients often emphasize the importance of other factors, such as dizziness, pain, fatigue, cognitive difficulties, and their satisfaction or regret with treatment decisions, as critical symptoms and experiences with healthcare delivery affecting their social and emotional well-being (9,10). This highlights the need for more robust tools and studies that prioritize the patient perspective, enabling healthcare professionals to deliver care that truly aligns with patient values and improves overall well-being.

The Penn Acoustic Neuroma Quality of Life-questionnaire (PANQOL) was developed in 2010 as a diseases-specific health-related quality of life (HrQoL) measure (11). It was translated into German by Kristin et al. in 2017 (12). Its domains focus primarily on physical and emotional aspects of QOL, such as hearing, balance, and facial function, but may not fully capture other important domains such as cognitive function, work ability, or treatment satisfaction (11). The German PANQOL questionnaire, although effective in measuring QOL, has several limitations. The “General Health” domain showed poor reliability, making it inadequate for assessing QOL, while the “Pain” domain, with only one item, lacks reliability and would benefit from additional items. Strong correlations between domains raise concerns about the validity and specificity of the constructs. There may also be an imbalance between clinically important and less important aspects of QOL that clinicians should consider when counseling patients. In addition, the study focused on patients with small tumors, so results may be different for larger VS (13).

As already highlighted, patient-centered domains are becoming increasingly important as patient-centered care emphasizes holistic outcomes. Therefore, the Vestibular Schwannoma Quality of Life Index (VSQOL) was developed by Carlson et al. (14) in 2022. This 40-item questionnaire goes beyond the assessment of typical VS symptoms such as hearing loss, dizziness, and pain to include other important domains such as cognition, well-being, ability to work, and treatment satisfaction areas that are essential for patient-centered care but often lacking in other disease-specific QoL instruments. In a preceding study (10), the VSQOL was translated into German and its content validity was demonstrated. Yet, to date the German version of the VSQOL has not undergone further psychometric testing. In this study, VS patients also highlighted limitations in the current degree of patient-centeredness of German healthcare delivery, e.g. in the lack of integration of medical and non-medical care, hindering comprehensive biopsychosocial treatment and thus negatively impacting long-term outcomes (10).

Therefore, the primary aim of this study is to psychometrically test the German version to confirm its reliability and validity as a QoL assessment tool for German-speaking VS patients. In addition, our secondary aim is to assess the experiences of patient-centered care in VS patients and to further explore how these experiences affect their overall well-being.

## Methods and Analysis

### Study Design

This study is designed as a multicenter, cross-sectional validation study to evaluate the psychometric properties of the German version of the VSQOL and to explore patient-centered care experiences among German-speaking patients with VS. The study will collect data from multiple clinical sites in Germany and Switzerland to ensure a diverse sample of VS patients.

### Patient and Public Involvement

Patients were involved in the preliminary phase of the study, particularly in the translation and establishment of content validity of the German VSQOL through cognitive interviews. Their feedback was instrumental in refining the questionnaire to ensure cultural relevance and comprehensibility. Throughout the main study, two patient representatives from *Vereinigung Akustikus Neurinom e*.*V*. (VAN), a national patient organization on VS, will be consulted on recruitment strategies and dissemination of results. Meetings will be held online every three months. We are aiming for level 5 participation - inclusion. Patient representatives are involved in decision-making, but do not have decision-making power (15). The study results will be shared with participants and patient advocacy groups to ensure they are informed of the findings.

### Sample

The sample will include adult patients diagnosed with VS who were treated with microsurgery, radiotherapy and/or wait-and-scan. Exclusion criteria are reduced cognitive capacity and inadequate German language skills. Patients with a diagnosis of neurofibromatosis 2 are also excluded, as this condition is associated with additional symptoms that are better reflected by another QoL instrument, such as the Neurofibromatosis 2 Impact on QoL (NFTI-QOL) (16). A minimum of 280 participants is required, as calculated according to the Consensus-based Standards for the selection of health Measurement Instruments (COSMIN) checklist (17), which suggests a sample size equivalent to at least seven times the number of items in the questionnaire or at least 100 participants.

### Recruitment Strategies

Participants will be recruited using a multifaceted approach, including an online call via mailing lists for members of the VAN, a national patient organization with VS, and on-site recruitment at various specialized departments. Specifically, recruitment will take place at the Departments of Neurosurgery and Radiation Oncology at the University Medical Center Hamburg-Eppendorf (UKE) as well as at the Department of Neurosurgery and Department of Radiation Oncology at the University Hospital Heidelberg in Germany, the Department of Neurosurgery at the University Hospital Halle (Saale) in Germany, and the Department of Otolaryngology, Head & Neck Surgery at the University Hospital Zurich in Switzerland. This strategy allows us to reach a diverse patient population that includes individuals who have undergone different types of treatment, including surgical, radiation, and conservative management approaches (wait-and-scan).

### Financial and other rewards to participants

A prize draw involving participants who have opted to provide their contact details will take place at the conclusion of data collection. Ten randomly selected participants will receive 15€ in gift cards. This prize will be drawn among the LimeSurvey participants.

### Data collection

In this study, demographic (e.g., age, gender, employment status) and clinical variables related to the VS (e.g., tumor size in Koos grade, date of treatment) will be assessed using selfadministered questions. Several validated instruments will be used to comprehensively assess various dimensions of HrQoL and patient experiences. The primary tool will be the German VSQOL. We will assess the convergent and discriminant validity of the VSQOL Index scores by calculating Pearson and Spearman rank correlations with other general and disease-specific instruments (see Table 1).

**Table 1.**
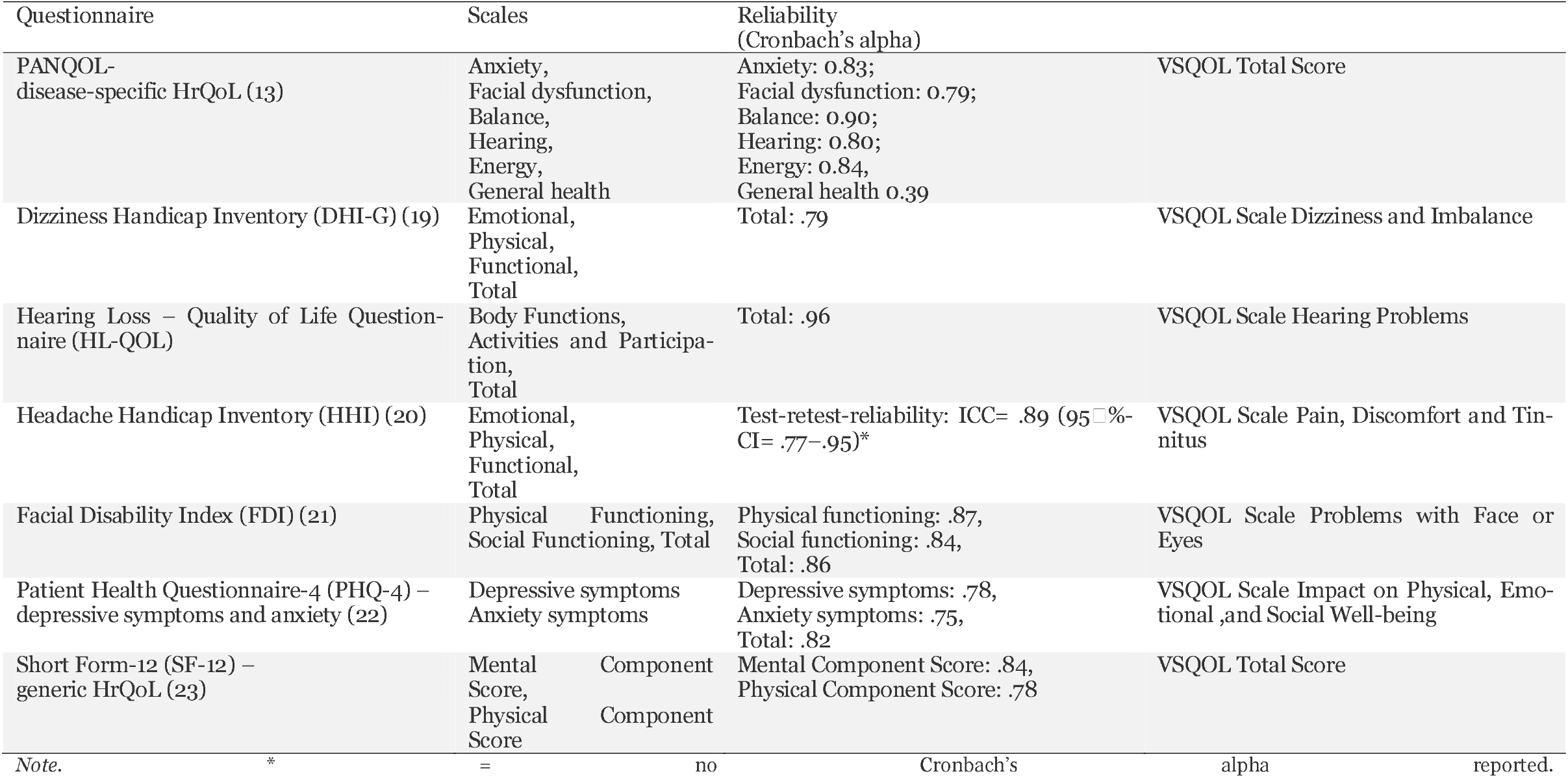
Questionnaires used for convergent validity.

| Questionnaire | Scales | Reliability<br>(Cronbach's alpha) |  |
| --- | --- | --- | --- |
| PANQOL-disease-specific HrQoL (13) | Anxiety,<br>Facial dysfunction,<br>Balance,<br>Hearing,<br>Energy,<br>General health | Anxiety: 0.83;<br>Facial dysfunction: 0.79;<br>Balance: 0.90;<br>Hearing: 0.80;<br>Energy: 0.84,<br>General health 0.39 | VSQOL Total Score |
| Dizziness Handicap Inventory (DHI-G) (19) | Emotional,<br>Physical,<br>Functional,<br>Total | Total: .79 | VSQOL Scale Dizziness and Imbalance |
| Hearing Loss – Quality of Life Question-<br>naire (HL-QOL) | Body Functions,<br>Activities and Participa-<br>tion,<br>Total | Total: .96 | VSQOL Scale Hearing Problems |
| Headache Handicap Inventory (HHI) (20) | Emotional,<br>Physical,<br>Functional,<br>Total | Test-retest-reliability: ICC= .89 (95%<br>CI= .77–.95)* | VSQOL Scale Pain, Discomfort and Tin-<br>nitus |
| Facial Disability Index (FDI) (21) | Physical Functioning,<br>Social Functioning, Total | Physical functioning: .87,<br>Social functioning: .84,<br>Total: .86 | VSQOL Scale Problems with Face or<br>Eyes |
| Patient Health Questionnaire-4 (PHQ-4) –<br>depressive symptoms and anxiety (22) | Depressive symptoms<br>Anxiety symptoms | Depressive symptoms: .78,<br>Anxiety symptoms: .75,<br>Total: .82 | VSQOL Scale Impact on Physical, Emo-<br>tional ,and Social Well-being |
| Short Form-12 (SF-12) –<br>generic HrQoL (23) | Mental Component<br>Score,<br>Physical Component<br>Score | Mental Component Score: .84,<br>Physical Component Score: .78 | VSQOL Total Score |

Note. \* = no Cronbach's alpha reported.

Additionally, the Experienced Patient-Centeredness Questionnaire (EPAT) (18) will evaluate patients’ experiences with patient-centered care (secondary objective). This tool offers a comprehensive framework for evaluating patient-centeredness, giving users the flexibility to assess each dimension using a four-item module or to select specific modules based on their unique needs. It comprises 16 distinct modules, 8 of which were chosen in collaboration with the VAN for this study. These selected dimensions encompass key aspects of patient-centered care: essential characteristics of the clinician, clinician–patient relationship, patient as a unique person, biopsychosocial perspective, clinician–patient communication, integration of medical and nonmedical care, coordination and continuity of care, and patient information. The EPAT demonstrated a bi-factorial structure, with McDonald’s Omega hierarchical indicating strong reliability for the general factor (outpatients: 0.868; inpatients: 0.908). In contrast, reliability for the specific dimensions varied considerably, ranging from 0.214 to 0.872 for outpatients and from 0.128 to 0.847 for inpatients (18).

## Data management

All participant data will be anonymized using unique identification codes to ensure privacy and entered into a secure, encrypted database with access restricted to authorized research personnel. Data integrity will be maintained through a standardized entry protocol with regular checks and audits to address discrepancies. Ongoing validation processes, including cross-referencing entries with source documents, will ensure data quality. Statistical analyses will be performed using SPSS (24), with results carefully documented for transparency and reproducibility.

Data storage will comply with ethical and regulatory standards, and, upon study completion, will be securely archived with limited access for the required retention period. A robust privacy policy will be enforced, including strict anonymization procedures, encrypted databases, and controlled access. The study will adhere to the Declaration of Helsinki (25) and relevant national regulations. LimeSurvey (26), a GDPR-compliant platform, will be used to collect and manage online survey responses securely, with encrypted transmissions and anonymized storage to enhance confidentiality. Data collection at the University Hospital Heidelberg is going to be performed using the in-house developed patient portal “Chili” in cooperation with the MIRO-Team and consecutive storage of the data in the research database of HIRO.

## Statistical Analyses

### Primary Objective: Psychometric testing

To ensure that the German version of the VSQOL is a reliable and valid instrument for assessing quality of life in patients with VS, a comprehensive psychometric validation will be conducted. Reliability will be assessed primarily through internal consistency, evaluated using Cronbach’s alpha. This statistic measures the degree to which items within a scale are interrelated, indicating the extent to which they collectively capture the same underlying construct. A Cronbach’s alpha of >□.80 will be considered excellent for the total score, while values above .70 will be deemed acceptable for the individual subscales. These thresholds align with established psychometric standards and will help confirm the internal coherence of the questionnaire across its various domains. In addition, test-retest reliability will be assessed by calculating intraclass correlation coefficients in a subset of patients after 14 days to determine the stability of VSQOL scores over time.

Construct validity will be addressed by confirmatory factor analysis, which will test whether the factor structure of the German VSQOL is consistent with the theoretical model established in the original English version. To account for differences between the recruitment samples, we will first conduct a detailed descriptive analysis of the subgroups (outpatient clinics and online samples). This analysis will include examining means, item difficulties and other item characteristics, following the same approach as for the full dataset. The aim is to identify any variations or trends specific to each subgroup and to ensure that the data from these different recruitment sources are thoroughly understood before proceeding to more complex analyses. In addition, we plan to assess measurement invariance by estimating the model from the CFA for the subgroups. However, given the complexity of the model and the potential for small sample sizes in the subgroups, there is a risk that the model may not converge.

Convergent validity will be evaluated by examining the correlations between domain scores of the VSQOL and those of established quality of life instruments. Strong positive correlations (e.g., r ≥ .90) will indicate a high degree of convergent validity, demonstrating that the VSQOL effectively captures constructs similar to those measured by validated tools in the field. This will ensure that the VSQOL captures related but distinct constructs as expected. We expect the VSQOL Index scores to show strong correlations with instruments measuring similar constructs, but low or no correlations with those measuring different or unrelated concepts. Specifically, positive correlations in the range of .5 to .8 are expected between the VSQOL scales and corresponding validated questionnaires.

To manage missing values, we will employ a systematic approach to ensure the integrity and validity of the analysis. Initially, the extent and pattern of missing data will be examined to determine whether it occurs randomly or systematically. For data deemed missing completely at random (MCAR), appropriate imputation methods such as multiple imputation or mean imputation will be used. If the missing data are not random, techniques such as full information maximum likelihood (FIML) estimation or sensitivity analyses will be applied to account for potential biases (27). Moreover, items or cases with excessive missingness (>70% of data) may be excluded from specific analyses if justified. All handling of missing data will be transparently documented to maintain methodological rigor.

### Secondary Objective: Exploration of Experienced Patient-Centeredness

For the secondary objective of evaluating the experience of patient-centeredness in VS patients, statistical analysis will focus on exploring how perceptions of patient-centeredness influence QoL. Descriptive statistics will also be used to summarize patient feedback on their care experience, providing a detailed view of the overall patient-centered care experience. Together, these analyses will provide valuable insights into how well care is aligned with patients’ needs and preferences and its impact on their overall well-being. We will use correlation analysis to examine relationships between scores on the EPAT and the German VSQOL. This analysis will help to determine the strength and direction of associations between patient-centeredness and QOL outcomes.

## 3. Discussion

This study will adhere to the highest ethical standards to ensure the protection and welfare of all participants. Ethical approval has already been obtained from the local ethics committee for the Hamburg site and the online study. Approval was also sought from the Institutional Review Boards at the other participating sites prior to study initiation. Participants will be thoroughly informed of the study objectives, procedures, potential risks and benefits through a comprehensive informed consent process. Informed consent will be obtained in writing from each participant to ensure voluntary participation and a clear understanding of their rights. Participants will be explicitly informed that they have the right to withdraw from the study at any time before their data are anonymized, without any consequences or impact on their medical care.

Results of the study will be disseminated through multiple channels to engage both the academic community and the general public. Results will be published in peer-reviewed journals and presented at relevant conferences, contributing to the scientific understanding of QoL in patients with VS. In addition, results will be shared with participating patients, healthcare professionals, and patient advocacy groups to inform clinical practice and improve patient care. Members of the VAN can voluntarily choose to receive a summary of the study results by email. This ensures that they are directly informed of the study results.

## Abbreviation Full Term

VS: Vestibular Schwannoma
QoL: Quality of Life
HrQoL: Health-related Quality of Life
PANQOL: Penn Acoustic Neuroma Quality of Life Questionnaire
VSQOL: Vestibular Schwannoma Quality of Life Index
VAN: Vereinigung Akustikus Neurinom e.V.
NFTI-QOL: Neurofibromatosis 2 Impact on Quality of Life
COSMIN: Consensus-based Standards for the Selection of Health Measurement Instruments
UKE: University Medical Center Hamburg-Eppendorf
EPAT: Experienced Patient-Centeredness Questionnaire
FDI: Facial Disability Index
HL-QOL: Hearing Loss – Quality of Life Questionnaire
DHI-G: Dizziness Handicap Inventory – German version
HHI: Headache Handicap Inventory
PHQ-4: Patient Health Questionnaire-4
SF-12: Short Form-12
SPSS: Statistical Package for the Social Sciences
GDPR: General Data Protection Regulation
MIRO: Medical Informatics and Research Organisation
HIRO: Heidelberg Institute for Research Organisation
MCAR: Missing Completely At Random
FIML: Full Information Maximum Likelihood
ICC: Intraclass Correlation Coefficient
CFA: Confirmatory Factor Analysis

## Ethics approval and consent to participate

Ethical approval for this study was obtained from the relevant institutional review boards at all participating sites, including the University Medical Center Hamburg-Eppendorf and collaborating centers in Halle (Saale), Heidelberg and Zurich. Prior to data collection, all participants will receive detailed written and verbal information about the study’s objectives, procedures, potential risks, and benefits. Informed consent will be obtained in writing from each participant, ensuring voluntary participation. Participants will be assured that they may withdraw from the study at any time without any consequences for their medical care. The study will be conducted in accordance with the Declaration of Helsinki and applicable national data protection regulations.

### Authors’ contributions

MR and MS conceptualized and planned the study. MR wrote the manuscript. LD, AB, JW, BB, AH, and IS will participate in the recruitment of participants. All authors reviewed and approved the final manuscript.

### Data availability statement

The datasets used and/or analysed during the current study are available from the corresponding author on reasonable request.

## Acknowledgements

We would like to thank the members of VAN for their support during all stages of this study.

